# Metabolic and volumetric alterations in the basal ganglia and the cerebellum in dopa-responsive dystonia in symptomatic and asymptomatic *GCH1* mutation carriers

**DOI:** 10.64898/2026.01.23.26344694

**Authors:** Jannik Prasuhn, Leon van Well, Marta M. Pokotylo, Feline Hamami, Joke-Lina Aßmann, Katja Lohmann, Maximilian G. Ködderitzsch-Mertins, Julia Henke, Jan Uter, Alexander Münchau, Christine Klein, Anne Weissbach, Norbert Brüggemann

## Abstract

**Background:** Dopa-responsive dystonia is caused by pathogenic variants in the *GCH1* gene. Although its clinical features and reduced penetrance are known, *in vivo* metabolic and structural alterations in symptomatic (sMC) and asymptomatic mutation carriers (aMC) remain poorly understood.

**Objectives:** To characterize volumetric and neurometabolic brain changes of *GCH1* mutation carriers and explore their relationship with clinical severity.

**Methods:** We studied 20 sMC, 5 aMC, and 25 mutation-free healthy controls (HC) using volumetric MRI combined with ^31^phosphorus magnetic resonance spectroscopy imaging (^31^P-MRSI) of the basal ganglia and cerebellum. ANCOVA was used for group comparisons, and correlations were assessed with clinical symptom severity rating scales.

**Results:** Volumetric analyses revealed enlarged globus pallidus (+16.6%, p = 0.001) and putamen (+7.2%, p = 0.031) volumes in carriers and increased cerebellar gray matter in aMC (+8.0%, p = 0.050). NAD levels were significantly reduced in the basal ganglia of carriers (NAD/Pi: -14.7%, p = 0.046; NAD/ATP-α: -15.5%, p = 0.018). In the cerebellum, aMC demonstrated elevated high-energy phosphate ratios ((ATP-α+PCr)/Pi: +23.7%, p = 0.017; ATP-α/Pi: +21.3%, p = 0.046; PCr/Pi: +25.2%, p = 0.009) compared with sMC and HC. Smaller cerebellar volumes correlated with greater dystonia severity (BFMDRS, ρ = -0.475, p = 0.019) while lower basal ganglia NAD ratios correlated with higher MDS-UPDRS-III (ρ = -0.472, p=0.041) and TWSTRS scores (ρ = -0.477, p = 0.039).

**Conclusions:** ^31^P-MRSI and volumetric MRI reveal region- and subgroup-specific metabolic and structural alterations in *GCH1* mutation carriers, linking basal ganglia vulnerability and cerebellar adaptation to clinical severity.

## INTRODUCTION

Dopa-responsive dystonia (DRD) represents a clinically and genetically heterogeneous group of dystonia-parkinsonism most frequently caused by autosomal-dominant inherited mutations in the *GCH1* gene (DYT/PARK-*GCH1*).[1] The disorder, also known as Segawa syndrome, is characterized by early-onset dystonia with marked diurnal fluctuations, additional Parkinsonian features in some patients, and a robust and lasting therapeutic response to low doses of levodopa.[2]

Mutations in *GCH1* impair the biosynthesis of tetrahydrobiopterin (BH4), an essential cofactor for tyrosine hydroxylase, the rate-limiting enzyme in dopamine synthesis. The resulting reduction in dopamine availability particularly affects the striatum and related basal ganglia circuits, crucial for controlling movement.[3,4] Post-mortem studies and biochemical analyses demonstrated reduced dopamine content in striatal tissue of affected individuals, yet *in vivo* markers reflecting these metabolic disturbances remain scarce.[3] Moreover, *GCH1* mutations show incomplete penetrance with some carriers developing dystonia or Parkinsonism (symptomatic mutation carriers, sMC), while others remain clinically unaffected (asymptomatic mutation carriers, aMC).[1] This variability is also present at the intrafamilial level, highlighting the importance of compensatory mechanisms that may mitigate the functional consequences of impaired dopamine synthesis.

There is increasing evidence that dystonia is associated not only with basal ganglia dysfunction but also with alterations in the cerebellum and cerebello-thalamo-striatal networks.[4–6] Neuroimaging and animal studies in isolated and combined dystonias have suggested that the cerebellum may exert a modulatory or partially compensatory role.[5,7] Whether similar mechanisms are present in DRD, and in particular in aMC, remains an open question.

Advanced neuroimaging offers a unique opportunity to investigate these issues *in vivo*. ^31^Phosphorus magnetic resonance spectroscopy imaging (^31^P-MRSI) allows the quantification of high-energy phosphate (HEP) metabolites such as alpha adenosine triphosphate (ATP-α) and phosphocreatine (PCr), as well as nicotinamide adenine dinucleotide (NAD), which are sensitive markers of cellular energy metabolism and mitochondrial function.[8] Previous work in other movement disorders demonstrated that alterations in NAD and HEP ratios reflect impaired bioenergetic capacity and may serve as early indicators of neurodegenerative or neurodevelopmental processes.[9–12] Structural magnetic resonance imaging (MRI) with automated volumetric segmentation provides robust measures of subcortical and cerebellar anatomy, primary involved in the dopaminergic motor circuits, allowing detection of subtle structural alterations that may reflect either neurodevelopmental or neurodegenerative changes or compensatory plasticity.[13]

Despite the well-characterized genetic and clinical profile of DRD, systematic multimodal imaging studies investigating neurometabolic and structural correlates in *GCH1* mutation carriers (MC) are lacking. In this study, we set out to investigate the neurometabolic and structural features of *GCH1*-associated DRD using a multimodal imaging approach, combining ^31^P-MRSI of the basal ganglia and cerebellum with volumetric MRI in a cohort of sMC, aMC, and age- and sex-matched, mutation-free healthy controls (HC). Our objectives were to determine whether alterations in NAD and HEP metabolism can be detected *in vivo*, to assess whether volumetric differences in basal ganglia and cerebellar structures accompany these metabolic features, and to explore the relationship between imaging markers and clinical features.

## METHODS

### Recruitment and clinical characterization

The study protocol was reviewed and approved by the Ethics Committee of the University of Lübeck, and all procedures complied with the Declaration of Helsinki. Written informed consent was obtained from every participant prior to enrollment. Participants were identified through specialized in- and outpatient clinics and from existing research cohorts. Individuals were classified as either MC or HC. MC were further separated into sMC and aMC subgroups based on the presence or absence of manifest motor features. All participants underwent a standardized baseline assessment that included medical history, demographic information, and screening for contraindications to MRI. Genetic status was confirmed by genetic testing (sequencing and screening for copy number variants). Current medication use and previous neurological diagnoses were documented. Neurological examinations were performed by experienced specialists and were video-documented and rated in a blind manner.[14,15] Motor impairment was rated using the motor part of the Movement Disorder Society Unified Parkinson’s Disease Rating Scale (MDS-UPDRS-III) [16], and dystonia severity was assessed using the Burke-Fahn-Marsden Dystonia Rating Scale (BFMDRS motor/disability) [17] and the Toronto Western Spasmodic Torticollis Rating Scale (TWSTRS)[18].

### Neuroimaging acquisition and analyses

All imaging was conducted on a 3 Tesla Siemens MAGNETOM Skyra scanner at the Center for Brain, Behavior, and Metabolism (CBBM) Core Facility. sMC were investigated in the Levodopa ON state. All datasets were reviewed by neuroradiologists to exclude previously undiagnosed structural abnormalities. Only complete scans were considered for subsequent analyses.

#### T1-weighted imaging

Structural data were acquired using a standardized three-dimensional (3D) magnetization-prepared rapid gradient-echo (MPRAGE) sequence, with total scan time of 5 minutes and 52 seconds. This protocol enabled isotropic imaging with coverage in sagittal, coronal, and axial planes, yielding sharp T1-weighted contrast through magnetization preparation and efficient acquisition with rapid gradient echoes. Parallel imaging was applied with an acceleration factor of two, reducing scan time and minimizing motion-related artifacts. Sequence parameters were: voxel size = 0.8 × 0.8 × 0.8 mm^3^, field of = 320 × 320 × 320 mm^3^, TR = 2300 ms, TE = 2.43 ms, TI = 1100 ms, and flip angle = 8°.

#### ^31^Phosphorus-magnetic resonance spectroscopy imaging

^31^P-MRSI was performed using a double-tuned quadrature head coil ( ^1^H/^31^P, RAPID Biomedical) and a three-dimensional chemical shift imaging free induction decay sequence. Acquisition parameters were voxel size = 30 × 30 × 30 mm^3^, field of view = 240 × 240 × 240 mm^3^, TR = 2000 ms, TE = 2.3 ms, flip angle = 50°, sixfold weighted averaging, bandwidth = 2000 Hz, vector size = 1024, and Hamming filter width = 100%. Broadband proton decoupling (WALTZ-4) was applied, while the nuclear Overhauser effect was disabled. Total scan time was 8 minutes and 4 seconds. Four voxels were placed in the basal ganglia and two in the cerebellar hemispheres. Placement was standardized using anatomical landmarks (e.g., anterior commissure and internal capsule for basal ganglia; vermis and fourth ventricle for cerebellum) to maximize coverage of gray matter and minimize contamination from adjacent structures (Supplementary Figure 1). Shimming was adjusted manually using a slightly larger shim volume than the predefined volume of interest

#### Neuroimaging postprocessing and quantification

Spectra were processed using a Siemens workstation with the Syngo MR ER11 software package, as in our previous studies.[9,19–21] Preprocessing steps included Fourier transformation, zero-filling, frequency and phase correction, curve fitting, and baseline adjustment. Ratios normalized to inorganic phosphate (Pi) were calculated for ATP-α, PCr, and the combined (ATP-α + PCr), with (ATP-α + PCr)/Pi defined as the primary outcome. ATP-α/Pi and PCr/Pi served as secondary variables. Additionally, we calculated NAD/Pi and NAD/ATP-α as primary outcome measures to assess the NAD metabolism. To ensure robustness, non-normalized levels were additionally analyzed in an exploratory fashion. For each participant, side-averaged values were obtained for the basal ganglia and cerebellum.

**Figure 1.**
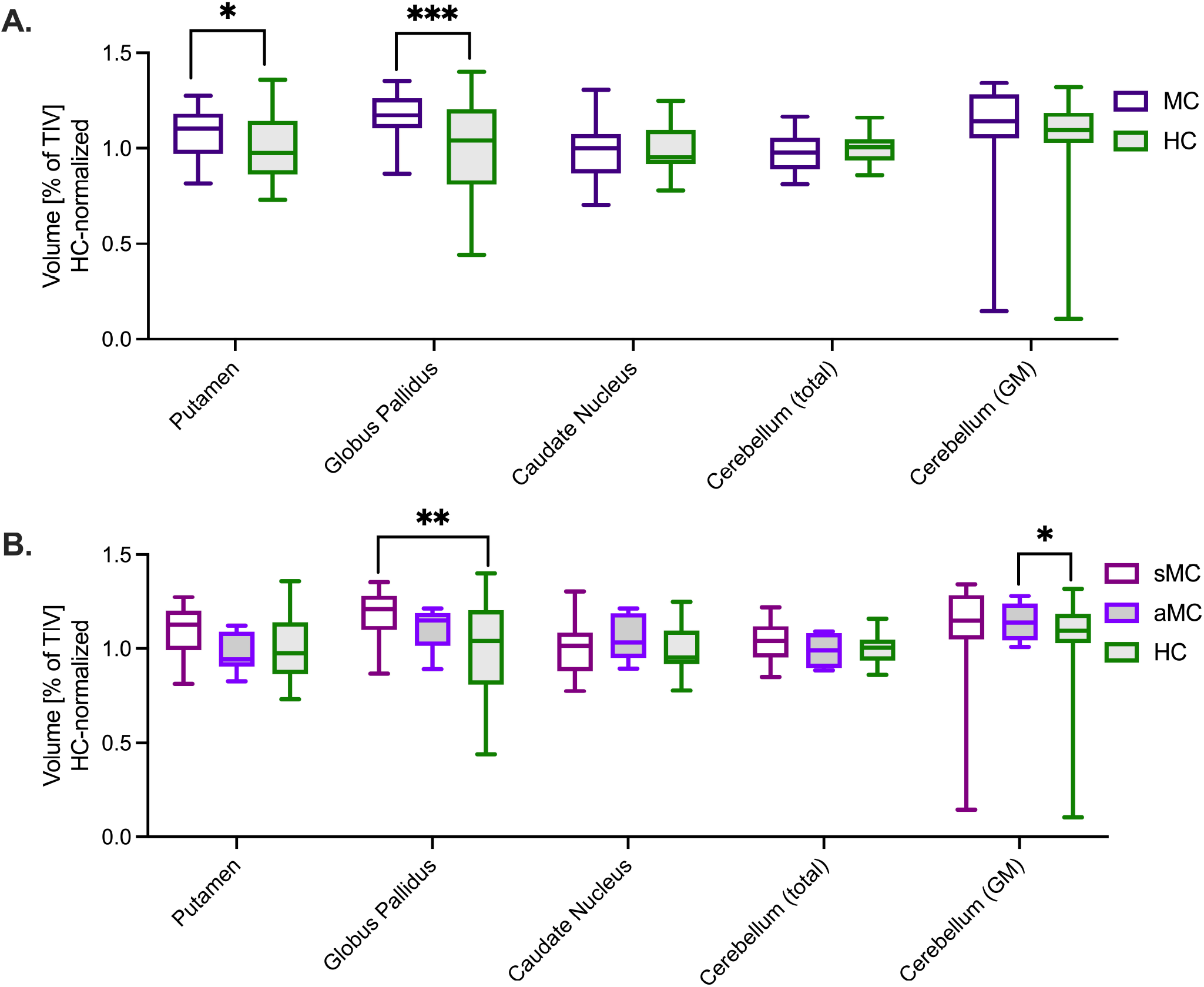
Group comparisons of subcortical and cerebellar volumetric measures across symptomatic and asymptomatic *GCH1* mutation carriers compared with mutation-free healthy controls. Brain volumes are presented as a percentage of total intracranial volume and normalized to the healthy control mean. In panel A, we present a comparison of mutation carriers (in purple) and healthy controls (in green). In panel B, we present a comparison of symptomatic mutation carriers (in pink), asymptomatic mutation carriers (in purple), and healthy controls (in green). The significant group differences are indicated by brackets (** < 0.01; * < 0.05). aMC = asymptomatic mutation carriers; GM = gray matter; HC = healthy controls; sMC = symptomatic mutation carries; TIV = total intracranial volume;

**Figure 2.**
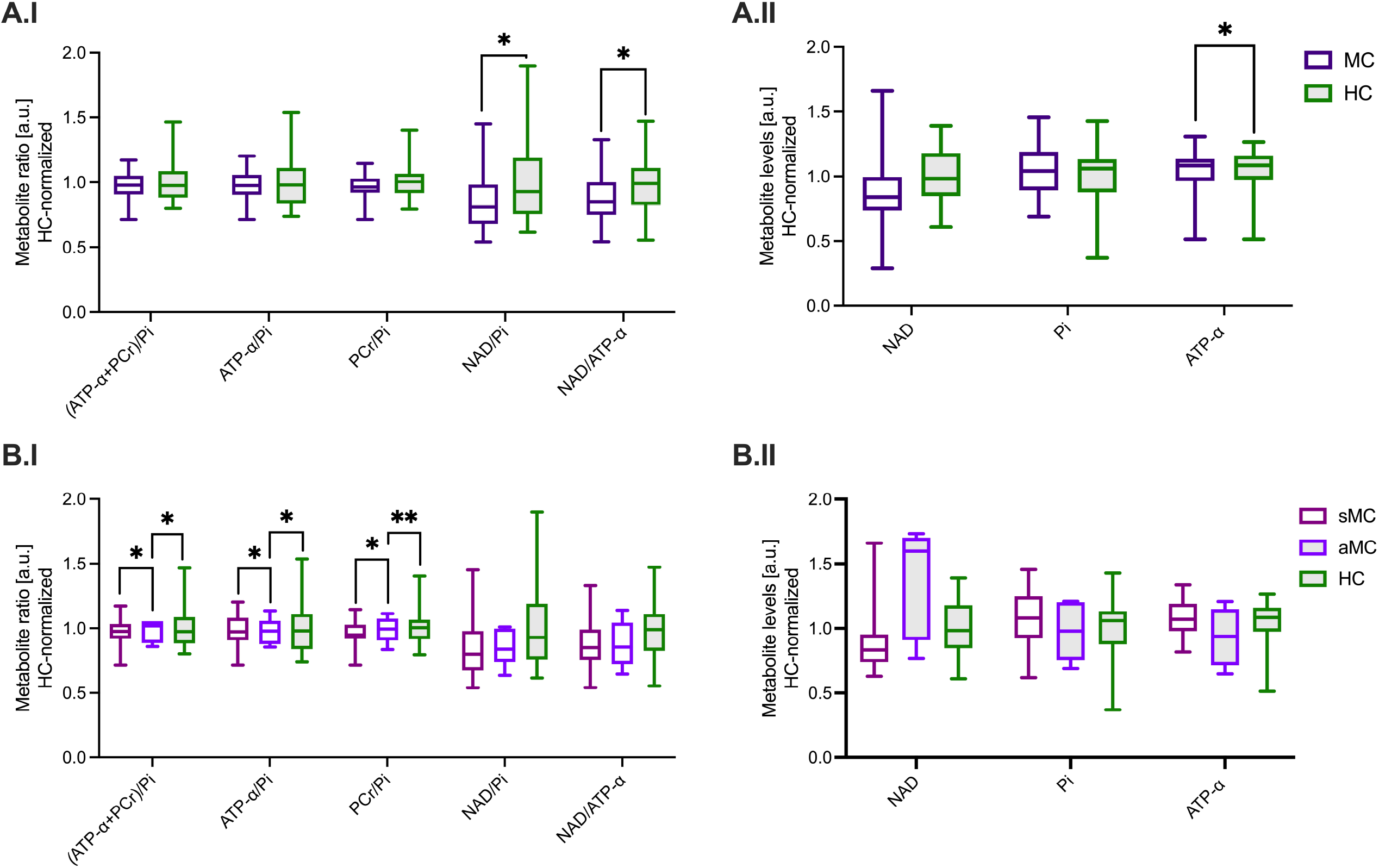
Group comparisons of ^31^phoshorus magnetic resonance spectroscopy-derived metabolite ratios in the basal ganglia across symptomatic and asymptomatic *GCH1* mutation carriers compared with mutation-free healthy controls. Demonstrated values are derived from the four voxels in the basal ganglia, expressed in arbitrary units and normalized to the healthy control mean. In panels A and B, we present the two-group and three-group derived ANCOVA results, respectively, comparing mutation carriers and healthy controls, as well as symptomatic and asymptomatic mutation carriers and healthy controls. In Panels A.I and B.I we present metabolite ratios and in panels A.II and B.II, the individual metabolites. Statistically significant group differences are indicated by brackets (* < 0.05). ATP-α = adenosine triphosphate phosphate alpha; aSMC = asymptomatic mutation carriers; a.u. = arbitrary units; HC = healthy controls; NAD = nicotinamide adenine dinucleotide; PCr = phosphocreatine; Pi = inorganic phosphate; sMC = symptomatic mutation carriers.

#### Volumetric analyses were performed using the volBrain platform

T1-weighted MPRAGE images were preprocessed through denoising, inhomogeneity correction, and spatial normalization to Montreal Neurology Institute (MNI) space. Total intracranial volume (TIV) was estimated, and tissue segmentation into gray matter (GM), white matter, and cerebrospinal fluid compartments was performed. Regional volumetry was derived from the volBrain submodules, providing automated parcellation of subcortical structures (caudate nucleus, putamen, globus pallidus) as well as total cerebellar volume and cerebellar GM.[22] All volumetric measures were quality-controlled through visual inspection of segmentation outputs. For group comparisons, TIV-normalized data were used to account for head size variability. Non-TIV-normalized volumetric values were retained for correlation analyses with ^31^P-MRSI-derived metabolite levels.

### Statistical analyses

All statistical procedures were conducted in Jamovi (version 2.3.21.0). Descriptive statistics summarize demographic and clinical variables across subgroups, reported as means and standard deviations. Distributional assumptions were examined using quantile-quantile (Q-Q) plots and formally tested using the Shapiro-Wilk test. Group comparisons of neuroimaging outcomes were performed using analysis of covariance (ANCOVA). Dependent variables included both normalized and non-normalized metabolite levels, as well as volumetric measures. The group assignment served as the fixed factor, while age, sex, and total TIV were included as covariates to account for potential confounding effects. Analyses were first carried out in a two-group design, contrasting all MC with HC, and subsequently extended to a three-group design that distinguished between symptomatic sMC, aMC, and HC. When ANCOVA yielded significant results, post hoc pairwise comparisons were performed using Bonferroni correction to control for type I error. Finally, correlation analyses were performed to explore associations between ^31^P-MRSI-derived metabolite values, volumetric measures of basal ganglia and cerebellar structures, and clinical severity scores (BFMDRS, TWSTRS, MDS-UPDRS-III). Spearman’s rank correlation coefficients were reported.

## RESULTS

### Demographics and clinical characteristics

The study included 50 participants: 20 sMC, 5 aMC, and 25 HC (see Table 1, Supplementary Table 1). Mean age was comparable across groups (sMC: 43.5 ± 17.3; aMC: 53.0 ± 25.7; and HC: 46.6 ± 19.6 years). In sMC, the average age at onset (AAO) was 10.7 ± 12.3 years, and the average levodopa equivalent daily dose (LEDD) was 282.8 ± 189.4 mg/d. Clinical scores in sMC in the ON state suggested a sufficient degree of symptom control with a BFMDRS mobility score of 3.9 ± 3.4, and a BFMDRS severity score of 4.3 ± 5.2, TWSTRS scores of 18.8 ± 10.6, and mild Parkinsonian symptoms with an MDS-UPDRS-III score of 8.5 ± 8.0.

**Table 1.**
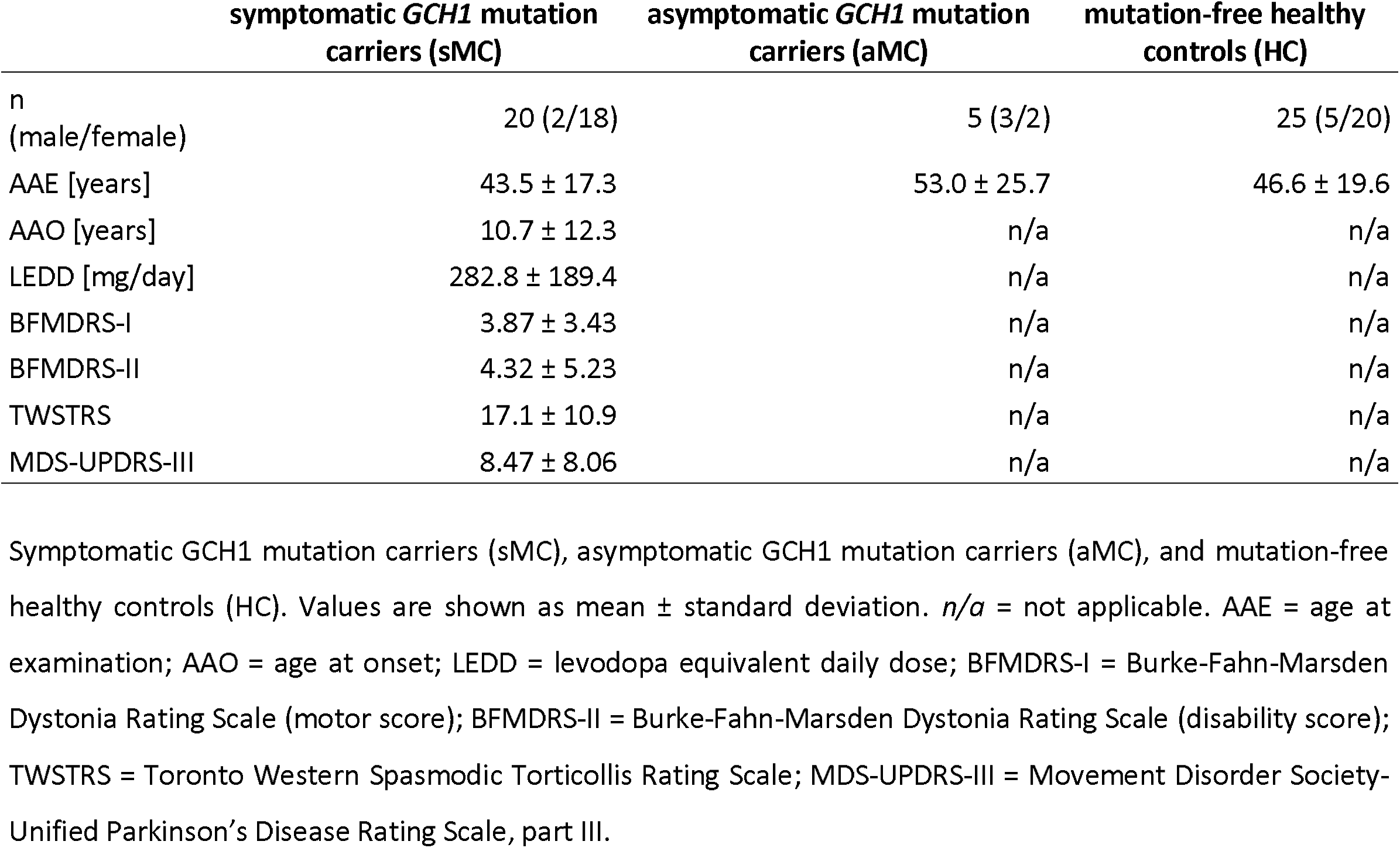
Demographic and clinical characteristics of symptomatic and asymptomatic *GCH1* mutation carriers compared with mutation-free healthy controls. Symptomatic GCH1 mutation carriers (sMC), asymptomatic GCH1 mutation carriers (aMC), and mutation-free healthy controls (HC). Values are shown as mean ± standard deviation. *n/a* = not applicable. AAE = age at examination; AAO = age at onset; LEDD = levodopa equivalent daily dose; BFMDRS-I = Burke-Fahn-Marsden Dystonia Rating Scale (motor score); BFMDRS-II = Burke-Fahn-Marsden Dystonia Rating Scale (disability score); TWSTRS = Toronto Western Spasmodic Torticollis Rating Scale; MDS-UPDRS-III = Movement Disorder Society-Unified Parkinson’s Disease Rating Scale, part III.

### Increased basal ganglia and cerebellar gray matter volumes in GCH1 mutation carriers

In the two-group ANOVA (MC vs. HC), basal ganglia volumes were higher in MC than in HC (see Figure 1). Putamen volume was 0.579 ± 0.073% of intracranial volume in MC vs. 0.540 ± 0.092% in HC (+7.2%; F(1,46) = 4.932, p = 0.031, η^2^ = 0.060). Globus pallidus volume was 0.197 ± 0.022% vs. 0.169 ± 0.044% (+16.6%; F(1,46) = 11.950, p = 0.001, η^2^ = 0.143). Caudate nucleus volumes did not differ (0.491 ± 0.072% vs. 0.500 ± 0.058%; -1.8%; F(1,46) = 0.556, p = 0.460, η^2^ = 0.008). The cerebellar volume was similar between MC and HC (9.773 ± 0.970% vs. 9.530 ± 0.706%; +2.5%; F(1,46) = 1.111, p = 0.297, η^2^ = 0.021), as well as cerebellar GM between MC and HC (7.986 ± 0.883% vs. 7.626 ± 0.551%; +4.7%; F(1,46) = 3.402, p = 0.072, η^2^ = 0.061).

In the three-group ANCOVA (sMC, aMC, HC), an increase in globus pallidus volume was most pronounced in sMC (see Figure 1). Volumes were 0.200 ± 0.022% in sMC, 0.188 ± 0.022% in aMC, and 0.169 ± 0.044% in HC (+18.3% and +11.2%; F(2,45) = 6.090, p = 0.005, η^2^ = 0.146; post hoc sMC vs. HC p = 0.004). Putamen volumes were 0.589 ± 0.070%, 0.540 ± 0.078%, and 0.540 ± 0.092% (+9.1% and 0.0%; F(2,45) = 2.571, p = 0.088, η^2^ = 0.064) but did not differ significantly after correction. Caudate nucleus volumes were similar across groups (0.501 ± 0.071%, 0.450 ± 0.065%, 0.500 ± 0.058%; F(2,45) = 0.963, p = 0.390, η^2^ = 0.027). Total cerebellar volume was 9.721 ± 0.964% in sMC, 9.983 ± 1.077% in aMC, and 9.530 ± 0.706% in HC (+2.0% and +4.8%; F(2,45) = 1.580, p = 0.216, η^2^ = 0.059). Cerebellar GM significantly differed between groups (7.923 ± 0.884%, 8.236 ± 0.928%, and 7.626 ± 0.551%; +3.9% and +8.0%; F(2,45) = 3.290, p = 0.046, η^2^ = 0.113; post hoc aMC vs. HC p = 0.050).

### Reduced nicotinamide adenine dinucleotide ratios in the basal ganglia of GCH1 mutation carriers

In the two-group ANCOVA (MC, HC), normalized ^31^P-MRSI ratios of HEPs did not differ significantly between MC and HC (see Figure 3). The combined (ATP-α + PCr)/Pi was 6.615 ± 0.681 in carriers and 6.821 ± 0.985 in HC (-3.0%; F(1,45) = 0.542, p = 0.465, η^2^ = 0.011). ATP-α/Pi was 3.112 ± 0.355 and 3.164 ± 0.578 (-1.6%; F(1,45) = 0.018, p = 0.783, η^2^ = 0.002), and PCr/Pi was 3.502 ± 0.374 and 3.657 ± 0.466 (-4.2%; F(1,45) = 1.521, p = 0.224, η^2^ = 0.030). NAD ratios were significantly reduced in carriers. NAD/Pi was 0.301 ± 0.071 vs. 0.353 ± 0.104 (-14.7%; F(1,45) = 4.199, p = 0.046, η^2^ = 0.084), and NAD/ATP-α was 0.097 ± 0.021 vs. 0.112 ± 0.025 (-15.5%; F(1,45) = 5.989, p = 0.018, η^2^ = 0.104). Non-normalized levels differed between groups, although with no statistical significance: NAD was 58.5 ± 16.0 vs. 63.6 ± 16.6 (-8.0%; p = 0.236), Pi was 195.9 ± 37.0 vs. 184.4 ± 35.9 (+6.2%; p = 0.147), and ATP-α was 603.1 ± 93.5 vs. 577.2 ± 113.1 (+4.5%; p = 0.095).

**Figure 3.**
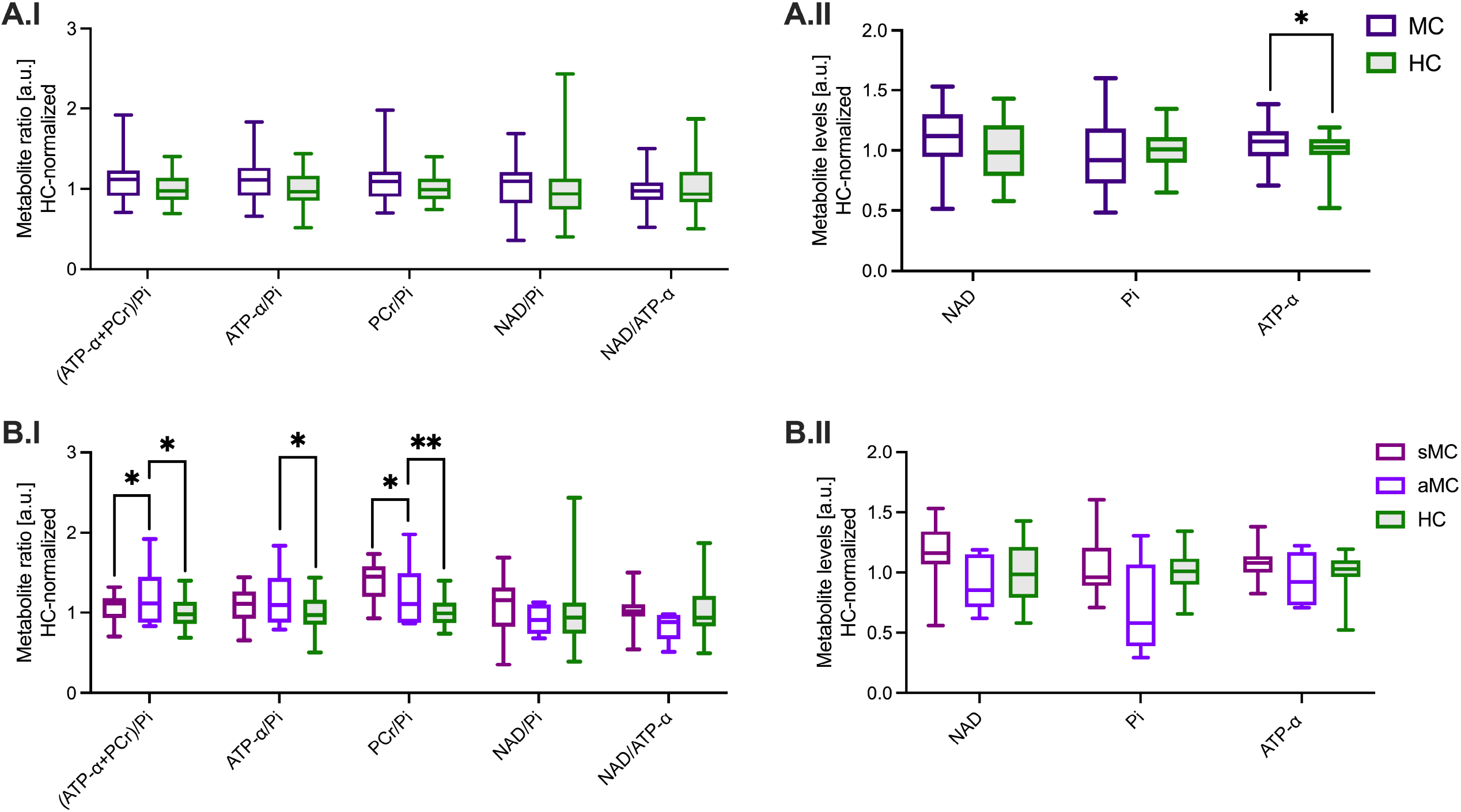
Group comparisons of ^31^phoshorus magnetic resonance spectroscopy-derived metabolite ratios in the cerebellum across symptomatic and asymptomatic *GCH1* mutation carriers compared with mutation-free healthy controls. Demonstrated values are derived from the two voxels in the cerebellum, expressed in arbitrary units and normalized to the healthy control mean. In panels A and B, we present the two-group and three-group derived ANCOVA results, respectively, comparing mutation carriers and healthy controls, as well as symptomatic and asymptomatic mutation carriers and healthy controls. In Panels A.I and B.I we present metabolite ratios and in panels A.II and B.II, the individual metabolites. Statistically significant group differences are indicated by brackets (** < 0.01; * < 0.05). ATP-α = adenosine triphosphate phosphate alpha; aSMC = asymptomatic mutation carriers; a.u. = arbitrary units; HC = healthy controls; NAD = nicotinamide adenine dinucleotide; PCr = phosphocreatine; Pi = inorganic phosphate; sMC = symptomatic mutation carriers.

In the three-group ANCOVA (sMC, aMC, HC), normalized HEP ratios showed no significant differences (see Figure 3). The combined (ATP-α + PCr)/Pi was 6.619 ± 0.711 in sMC, 6.599 ± 0.616 in aMC, and 6.821 ± 0.985 in HC (-3.0% and -3.3%; F(2,44) = 0.267, p = 0.767, η^2^ = 0.011). ATP-α/Pi was 3.132 ± 0.363, 3.034 ± 0.350, and 3.164 ± 0.578 (-1.0% and -4.1%; F(2,44) = 0.042, p = 0.959, η^2^ = 0.002). PCr/Pi was 3.486 ± 0.381, 3.565 ± 0.378, and 3.657 ± 0.466 (-4.7% and -2.5%; F(2,44) = 0.770, p = 0.469, η^2^ = 0.031). NAD ratios were lower in MC; however, with no statistical significance, NAD/Pi was 0.297 ± 0.075 in sMC, 0.317 ± 0.056 in aMC, and 0.353 ± 0.104 in HC (-15.9% and -10.2%; F(2,44) = 2.072, p = 0.138, η^2^ = 0.085). NAD/ATP-α was 0.095 ± 0.021, 0.106 ± 0.022, and 0.112 ± 0.025 (-15.2% and -5.4%; F(2,44) = 2.953, p = 0.063, η^2^ = 0.106). Non-normalized levels did not differ significantly, demonstrating numerically similar differences. NAD was 58.7 ± 15.5 in sMC, 57.6 ± 20.1 in aMC, and 63.6 ± 16.6 in HC (-7.7% and -9.4%; F(2,44) = 0.952, p = 0.394, η^2^ = 0.037). ATP-α was 619.6 ± 75.8, 536.8 ± 135.1, and 577.2 ± 113.1 (+7.4% and -7.0%; F(2,44) = 1.540, p = 0.226, η^2^ = 0.039). Pi was 200.5 ± 35.3, 177.6 ± 42.2, and 184.4 ± 35.9 (+8.8% and -3.7%; F(2,44) = 1.112, p = 0.338, η^2^ = 0.338).

### Increased high-energy phosphate ratios in the cerebellum of asymptomatic GCH1 mutation carriers

In the two-group ANCOVA (MC, HC), normalized ^31^P-MRSI ratios of HEPs were slightly higher in MC, with no statistical significance (see Figure 4). The combined (ATP-α + PCr)/Pi was 8.230 ± 1.913 in MC and 7.543 ± 1.475 in HC (+9.1%; F(1,45) = 1.276, p = 0.265, η^2^ = 0.026). ATP-α/Pi was 3.516 ± 0.869 vs. 3.218 ± 0.725 (+9.3%; F(1,45) = 1.036, p = 0.314, η^2^ = 0.021), and PCr/Pi was 4.714 ± 1.090 vs. 4.326 ± 0.789 (+9.0%; F(1,45) = 1.411, p = 0.241, η^2^ = 0.029). NAD ratios did not differ between groups: NAD/Pi was 0.379 ± 0.106 vs. 0.364 ± 0.152 (+4.1%; F(1,45) = 0.019, p = 0.890, η^2^ ≈ 0.000), and NAD/ATP-α was 0.109 ± 0.023 vs. 0.112 ± 0.031 (-2.7%; F(1,45) = 0.761, p = 0.388, η^2^ = 0.013). For non-normalized levels, ATP-α was higher in MC (583.5 ± 88.4) compared with HC (551.6 ± 83.0; +5.8%; F(1,45) = 4.056, p = 0.050, η^2^ = 0.055), whereas Pi (175.3 ± 47.9 vs. 176.7 ± 32.6; -0.8%; F(1,45) = 0.033, p = 0.856, η^2^ = 0.001) and NAD (63.7 ± 16.3 vs. 60.9 ± 15.8; +4.6%; F(1,45) = 0.108, p = 0.744, η^2^ ≈ 0.000) did not differ significantly.

**Figure 4.**
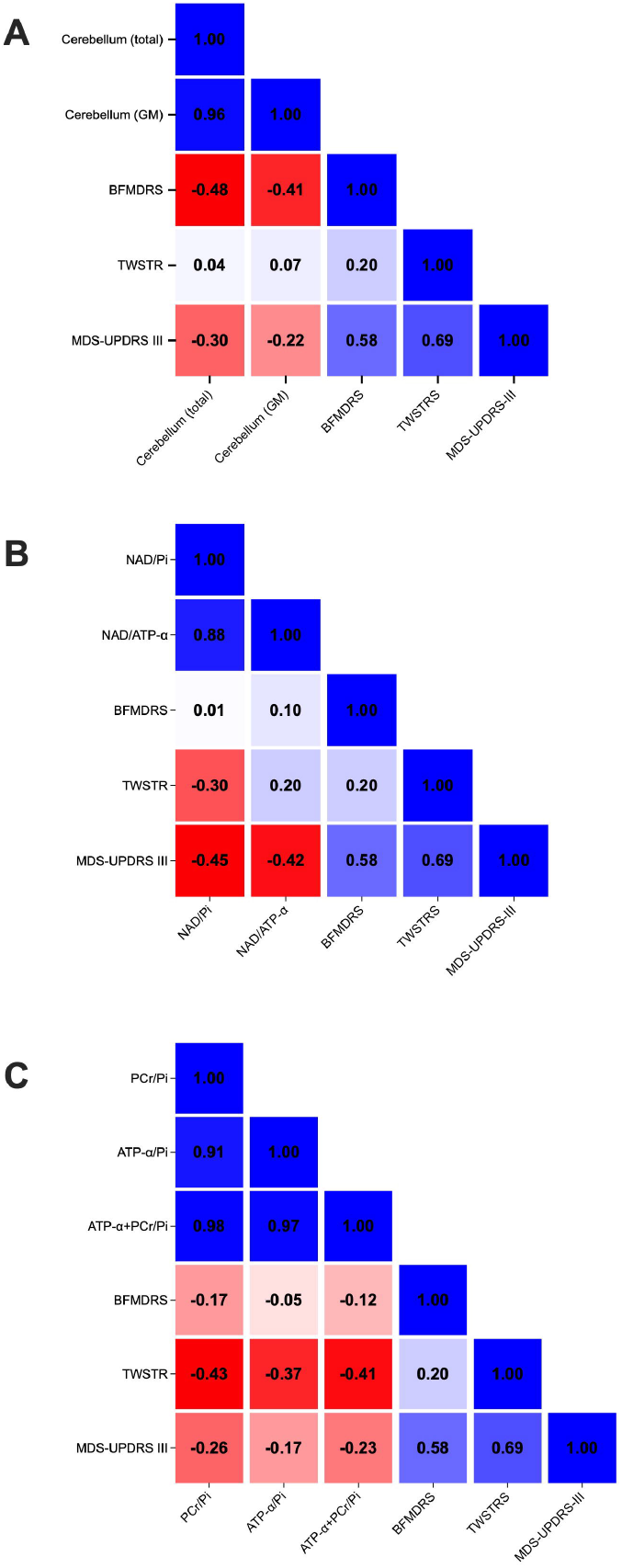
Correlation analyses of ^31^phosphorus magnetic resonance spectroscopy imaging metabolite ratios, volumetric measures, and clinical scores in *GCH1* mutation carriers. Demonstrated heatmaps present pairwise correlation coefficients (r) between ^31^phosphorus magnetic resonance spectroscopy imaging-derived measures and clinical scores. The presented heatmaps demonstrate the correlations between brain structure volumes (A), metabolite ratios in the basal ganglia (B) and cerebellum (C), and clinical scores. ATP-α = adenosine triphosphate alpha; BFMDRS = Burke-Fahn-Marsden Dystonia Rating Scale; GM = gray matter; MDS-UPDRS-III = Movement Disorder Society Unified Parkinson’s Disease Rating Scale III; NAD = nicotinamide adenine nucleotide; PCr = phosphocreatine; Pi = inorganic phosphate; TWSTRS = Toronto Western Spasmodic Torticollis Rating Scale.

In the three-group ANCOVA (sMC, aMC, HC), normalized ratios of HEPs revealed marked increases in aMC compared with sMC and HC (see Figure 4). (ATP-α + PCr)/Pi was 7.817 ± 1.427 in sMC, 9.880 ± 2.834 in aMC, and 7.543 ± 1.475 in HC (+3.6% and +30.9%; F(2,44) = 4.456, p = 0.017, η^2^ = 0.154; post hoc aMC vs. HC p = 0.016, aMC vs. sMC p = 0.027). ATP-α/Pi was 3.370 ± 0.759, 4.090 ± 1.134, and 3.218 ± 0.725 (+4.7% and +27.1%; F(2,44) = 3.314, p = 0.046, η^2^ = 0.117; post hoc aMC vs. HC p = 0.044). PCr/Pi was 4.445 ± 0.694, 5.791 ± 1.739, and 4.326 ± 0.789 (+2.7% and +33.9%; F(2,44) = 5.240, p = 0.009, η^2^ = 0.179; post hoc aMC vs. HC p = 0.008, aMC vs. sMC p = 0.014). NAD ratios remained unaltered. NAD/iP was 0.379 ± 0.117 in sMC, 0.382 ± 0.051 in aMC, and 0.364 ± 0.152 in HC (+4.1% and +5.0%; F(2,44) = 0.482, p = 0.621, η^2^ = 0.015). NAD/ATP-α was 0.111 ± 0.022, 0.099 ± 0.026, and 0.112 ± 0.031 (-0.9% and -11.6%; F(2,44) = 0.417, p = 0.662, η^2^ = 0.015). Non-normalized levels pointed in the same direction. ATP-α was 599.0 ± 73.1 in sMC, 521.4 ± 124.6 in aMC, and 551.6 ± 83.0 in HC (+8.6% and - 5.5%; F(2,44) = 2.114, p = 0.133, η^2^ = 0.060). Pi was 185.0 ± 41.1, 136.6 ± 58.0, and 176.7 ± 32.6 (+4.7% and -22.7%; F(2,44) = 2.721, p = 0.077, η^2^ = 0.091). NAD was 66.8 ± 14.9, 50.9 ± 16.8, and 60.9 ± 15.8 (+9.7% and -16.4%; F(2,44) = 0.289, p = 0.751, η^2^ = 0.009).

### Lower NAD ratios and reduced cerebellar volumes are associated with greater clinical severity in symptomatic GCH1 mutation carriers

Correlation analyses between ^31^P-MRSI ratios and clinical scores revealed that lower NAD ratios in the basal ganglia were associated with more severe motor impairment (see Figure 5). NAD/Pi correlated negatively with MDS-UPDRS-III scores (ρ = -0.472, p = 0.041), while NAD/ATP-α correlated negatively with TWSTRS scores (ρ = -0.477, p = 0.039) and showed a trend with MDS-UPDRS-III (ρ = -0.422, p = 0.072). Additionally, MDS-UPDRS-III scores correlated positively with both BFMDRS (disability score; ρ = 0.491, p = 0.033) and TWSTRS (ρ = 0.595, p = 0.007). Volumetric measures also showed disease-relevant associations. Smaller total cerebellar volume correlated with higher BFMDRS scores (ρ = -0.475, p = 0.019), and smaller cerebellar GM volume correlated similarly with higher BFMDRS (ρ = -0.408, p = 0.048) and vice versa.

## DISCUSSION

In this study, we combined ^31^P-MRSI and volumetric MRI to characterize neurometabolic and structural features of *GCH1*-related DRD *in vivo*. Three main observations emerged: (i) NAD ratios were consistently reduced in the basal ganglia of sMC and aMC, (ii) HEP ratios were increased in the cerebellum of aMC, and (iii) basal ganglia and cerebellar GM volumes were larger in aMC and sMC compared to HC. Importantly, lower basal ganglia NAD ratios and reduced cerebellar volumes were associated with greater clinical severity, linking imaging-derived markers to the phenotype. Our data support the concept of cerebellar compensation in DRD. The increase in cerebellar HEP metabolism, particularly in aMC, may reflect enhanced mitochondrial activity, counterbalancing basal ganglia dysfunction. These findings are supported by prior work demonstrating that the cerebellum contributes to the pathophysiology of dystonia through altered cerebello-thalamo-striatal signaling.[4,19] In our cohort, increased cerebellar GM volume in aMC further suggests structural plasticity, potentially underlying the absence of overt symptoms in this subgroup.

The reduced NAD ratios in the basal ganglia are consistent with a functional bioenergetic deficit. As NAD is a cofactor in the regeneration of BH4, reduced NAD availability may reflect impaired or exhausted BH4 recycling, further exacerbating the dopaminergic deficit caused by the pathogenic *GCH1* variants.[3,23] Enlargement of the globus pallidus and putamen in MC could reflect a compensatory response within striatal-pallidal circuits.[24–26] These findings need to be interpreted within the broader context of basal ganglia and cerebello⍰thalamo⍰cortical alterations that have been described across several forms of monogenic dystonia, including aMC of pathogenic *TOR1A* or *THAP1* variants. Structural and diffusion⍰based imaging studies in these conditions have shown abnormalities within striatal-pallidal circuits and in their connectivity with cerebellar and thalamo⍰cortical pathways, typically reflecting network dysfunction and reorganization rather than pronounced striatal atrophy. Supporting the network model, enlargement of the globus pallidus and putamen in MC may therefore represent a compensatory remodeling of striatal-pallidal loops in response to chronic dopaminergic dysfunction, which would be consistent with the absence of striatal atrophy in patients with DRD and distinguishes it from neurodegenerative dystonia⍰parkinsonism disorders, where progressive basal ganglia volume loss is prominent.[4,27– 31] Together, these results suggest that *GCH1*-related dystonia is characterized by a combination of basal ganglia vulnerability and cerebellar adaptation, with correlations between imaging markers and clinical severity adding further clinical relevance. Lower basal ganglia NAD ratios correlated with higher motor severity, while smaller cerebellar volumes were associated with more pronounced dystonic symptoms. Interestingly, higher cerebellar HEP ratios were observed in aMC, raising the possibility of a protective metabolic mechanism that may delay or prevent symptom manifestation.

Several limitations need to be considered. The cross-sectional design precludes conclusions about the temporal evolution of NAD and HEP metabolism in MC, necessitating longitudinal studies to determine whether reduced basal ganglia NAD ratios or elevated cerebellar HEP ratios precede, parallel, or follow clinical symptom manifestation, and whether they predict future progression or resilience. Additionally, the spatial resolution of ^31^P-MRSI was limited by the relatively large voxel size, potentially introducing partial volume effects, particularly in small subcortical nuclei, and potentially obscuring finer-grained regional differences. Furthermore, the spectral resolution did not allow for the differentiation between different oxidation states of NAD.[32,33] While our cohort was relatively large for a rare disease, subgroup analyses, especially in aMC, remain constrained by small sample sizes, limiting the statistical power to detect subtle effects. Moreover, sMC were examined in the ON medication state, raising the possibility that some of the observed effects may reflect treatment-related modulation rather than the disease process itself, as prior work demonstrates that levodopa administration can alter cerebral HEP metabolism.[20] OFF-state measurements could help disentangle disease-intrinsic from therapy-related changes, but may be challenging in patients with mobile dystonia; however, in some patients, if performed in the morning, they are generally feasible. Finally, ^31^P-MRSI cannot disentangle cellular sources of the metabolic signals, as well as the relative contribution of neuronal versus glial compartments, or of mitochondrial versus glycolytic ATP production. Similarly, volumetric MRI captures macrostructural changes but does not directly inform on microstructural adaptations that may underlie the observed enlargements.

In summary, our findings provide novel *in vivo* evidence that *GCH1*-related DRD is characterized by reduced basal ganglia NAD ratios, increased cerebellar high-energy phosphate metabolism, and structural adaptations in both basal ganglia and cerebellum. These results support a model in which cerebellar mechanisms compensate for basal ganglia dysfunction and highlight imaging-derived bioenergetic markers as promising tools for understanding variability in clinical expression. Longitudinal multimodal studies are warranted to clarify the temporal dynamics of these changes and their potential role as biomarkers for disease stratification and therapeutic monitoring.

## ACKNOWLEDGMENT

We sincerely thank all patients and study participants for their time, commitment, and invaluable contribution to this research. Their willingness to participate made this work possible and is deeply appreciated.

## AUTHOR CONTRIBUTION

**JP:** study conception and design, project supervision, data interpretation, manuscript drafting. **LvW:** data acquisition, data analysis, manuscript revision. **MMP:** data acquisition, formal analysis, visualization, manuscript revision. **FH:** data acquisition, data analysis, manuscript revision. **JLA:** data acquisition, data curation, manuscript revision. **KL:** (genetic) data acquisition and interpretation, manuscript revision. **MGKM:** methodology development, data analysis, manuscript revision. JH: data acquisition, data interpretation, manuscript revision. **JU:** data curation, statistical analysis, manuscript revision. **AM:** clinical interpretation, manuscript revision. **CK:** conceptual discussion, clinical interpretation, manuscript revision. **AW:** patient recruitment, clinical phenotyping, manuscript revision. **NB:** study conception, clinical supervision, data interpretation, manuscript revision.

## DATA AVAILABILITY

The data that supports the findings of this study will be made available upon reasonable request from the corresponding author.

